# Effects of acute psychological stress on blood cell-free mitochondrial DNA (cf-mtDNA): A crossover experimental study

**DOI:** 10.1101/2025.04.08.25325479

**Authors:** Caroline Trumpff, David Shire, Seonjoo Lee, Katie Stanko, Annette Wilson, Brett A. Kaufman, Martin Picard, Anna L. Marsland

## Abstract

In response to acute stress, prior studies have found an increase in circulating cell-free mitochondrial DNA (cf-mtDNA) and pro-inflammatory cytokines, highlighting two potential inter-related mechanisms by which stressors can get under the skin. However, prior studies lacked a resting control condition to isolate the effect of psychological stress from other aspects related to laboratory procedures. Here, we conducted a crossover experimental trial examining responses to a socio-evaluative stressor under laboratory conditions. 72 volunteers (age 20-50, 48% women) were tested on two occasions, counterbalanced, separated by at least a month. On one occasion, they were exposed to a 5-min socio-evaluative stressor (speech task), and on the other occasion, rested for the same period. Blood samples were obtained at 10 timepoints from pre- to 2 hours post-exposure to assess neuroendocrine (cortisol, catecholamines), pro-inflammatory cytokine (IL-6, IL-10, TNF-ɑ), and both plasma and serum cf-mtDNA responses.

Compared to the control visit, the stressor significantly increased anxiety, heart rate, blood pressure, cortisol, and norepinephrine (*p*’s<0.05-0.0001), confirming the psychobiological impact of the stressor. Unexpectedly, IL-6 and plasma cf-mtDNA increased (time effect *p*<0.0001) in both the stress and control conditions. While no significant effect of time was found for serum cf-mtDNA, plasma cf-mtDNA showed a bi-phasic response with an initial 22-24% increase at 5-10 min (g=0.07, stress-control visits), followed by a decrease and another 70-81% increase from 45 to 75 min (g=0.59 (stress visit), g=0.41 (control visit)). There were no significant associations between the pro-inflammatory and cf-mtDNA responses, pointing to their independent regulation. While mood, cardiovascular, and neuroendocrine reactivity were selectively induced by socio-evaluative stress, IL-6 and blood cf-mtDNA increased across both the stress and control conditions, suggesting that these biomarkers may reflect non-specific responses to the laboratory protocols (e.g., blood draw) rather than to socio-evaluative stress itself.

## 1. Introduction

Considerable evidence indicates that acute laboratory stressors elicit increases in circulating markers of inflammation ^1^. Individual differences in the magnitude of this response may reflect underlying characteristics influencing disease susceptibility ^2^. Further, the mechanisms linking psychological stress to inflammation remain partly unclear. Acute psychological stress activates multiple physiological systems leading to an increase in energy demand provided by mitochondria ^3^. Multiple aspects of mitochondria biology are affected by chronic and acute stress exposures ^4-6^ (reviewed in ^7^). Emerging evidence, including our own, suggests that brief laboratory stressors designed to model everyday stressors lead to elevated plasma and serum levels of cell-free mitochondrial DNA (cf-mtDNA) ^8-10^, a mitochondrial signaling marker associated with numerous disease states ^11^. In our previous work, exposure to a brief public speaking stressor resulted in a 1–2-fold increase in serum cf-mtDNA among healthy adults ^9^. This finding aligns with two other independent studies in which the Trier Social Stress Test led to a 60-70% increase in plasma cf-mtDNA ^8,10^. Some studies ^12-14^ (for a review, see ^11^), but not all ^15^, found an association between cf-mtDNA levels and inflammatory processes, suggesting cf-mtDNA could be hypothesized to be a mediator of stress-induced inflammation.

The initial studies showing acute cf-mtDNA reactivity to psychological stress ^8,9^ did not include a control condition, and many factors could have influenced the observed elevation in circulating levels of cf-mtDNA, including the presence of an intravenous catheter, the passage of time, and the stress of attending a laboratory session. In the current study, we conducted a well-controlled clinical trial to better characterize stress-induced regulation of cf-mtDNA and to examine its association with the increase in circulating proinflammatory mediators that is known to follow acute psychological stress and thought to contribute to health risk.

The mechanisms underlying the increase in cf-mtDNA following acute psychological stress exposure remain unclear. In our previous work, we explored predictors of stress-induced serum cf-mtDNA response and found that larger cf-mtDNA elevation after acute laboratory stress was associated with a higher pre-task heart rate and greater pre-to-post-task decreases in heart rate variability, a putative marker of parasympathetic nervous system activity, as well as greater pre-to-post-task increases in fatigue ^16^. Another study reported no association between stress-related plasma cf-mtDNA and stress-induced changes in cortisol or catecholamines, though these findings remain to be confirmed ^8^.

Here, we conducted the first controlled trial to examine blood cf-mtDNA responses to a brief laboratory stressor and their associations with psychological, cardiovascular, neuroendocrine, and pro-inflammatory cytokines reactivity. In this crossover experimental trial, participants attended two counterbalanced laboratory sessions. On one occasion, they were exposed to a 5-minute socio-evaluative public speaking stressor under laboratory conditions. On the other occasion, they rested quietly for the same period with identical assessment in the absence of the stressor. Given the different characteristics and differences in cf-mtDNA levels between plasma and serum ^17-20^, both biofluids were analyzed. We hypothesized that cf-mtDNA levels in both plasma and serum would increase within 30 minutes following the socio-evaluative stress task but remain unchanged in the control condition. We also hypothesized that stress-induced increases in cf-mtDNA would relate to concomitant increases in circulating proinflammatory mediators. Finally, exploratory analyses were conducted to explore predictors of the magnitude of cf-mtDNA response to acute psychological stress.

## Results

### 1.1. Acute psychological stress selectively induces mood and cardiovascular reactivity

Participants were 72 healthy volunteers from the *Mitochondria and Psychological Stress (MaPS)* study (48% female, mean age 32 years, see Table 1 for the descriptive characteristics). The within-subject protocol included two counterbalanced laboratory sessions: (1) a standardized 5-minute socio-evaluative public speaking task, and (2) a control condition ^9,21^ (Fig. 1A). At both sessions, blood samples were collected 5 minutes before and 5, 10, 20, 30, 45, 60, 75, 90 and 120 minutes after the onset of the task. 72 participants completed the first visit (n=37 stress visits) and 52 participants completed a second visit (n=26 stress visits).

**Figure 1.**
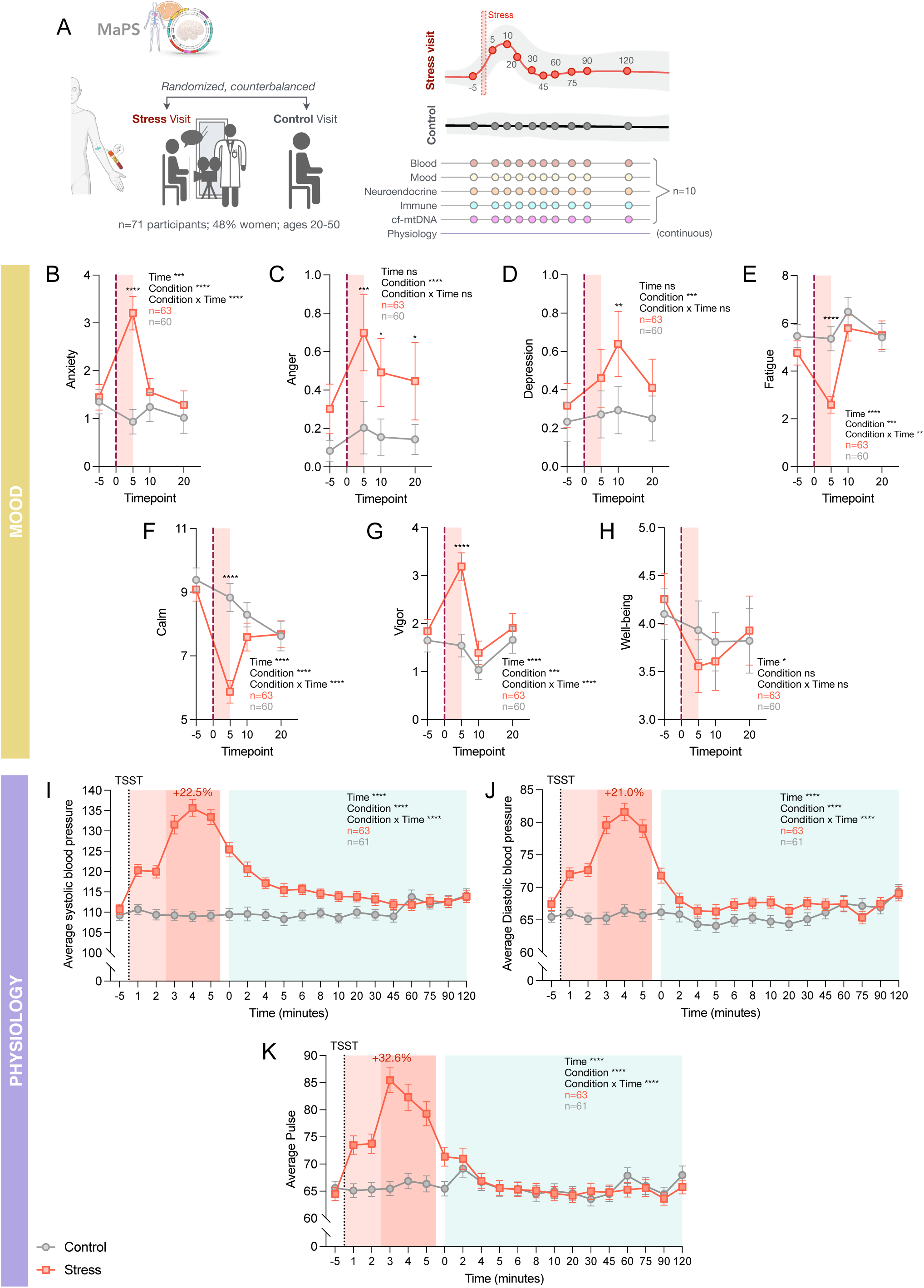
Mood and cardiovascular trajectories in response to acute psychological stress. **(A)** Overview of experimental design. Study participants attended a ‘stress visit’, where they were exposed to a modified version of the Trier Social Stress Test (TSST) speech task, and a ‘control visit’, during which they sat quietly for the same period as in the stress visit. Blood was collected at ten time points during each visit via intravenous catheter, and was analyzed to measure catecholamines, cytokines, and cell-free mitochondrial DNA. Cardiovascular measures and changes in self-reported mood were also recorded over the course of each visit. **(B-H)** Time courses of participants mood ratings in the stress and control conditions. Asterisks over symbols indicate a significant difference between control and stress values at the corresponding time point. **(I-K)** Time courses of participants cardiovascular measures in the stress and control conditions. Percentages indicate the maximum change between the time period occupied by the stress task (0 to +5 minutes) and the ‘-5’ time point. The two shades of red represent the preparation and speech phases of the stress task (0-2 minutes and 3-5 minutes, respectively). (B-K) Data shown as average ± standard error of the mean (SEM). Effect sizes and p-values from (B-K) mixed-effects analyses. *p<0.05, **p<0.01, ***p<0.001, ****p<0.0001.

As expected, self-reported mood differed across conditions, with increases in anxiety (condition x time *p* < 0.0001, Fig. 1B), decreases in fatigue and calmness (condition x time *p* < 0.01, Fig. 1E,F), and an increase in vigor (condition x time *p* < 0.0001, Fig. 1G) in response to the stress task when compared with the control condition. Interestingly, well-being decreased in both conditions (Fig. 1H). The stress task also selectively induced the expected cardiovascular response with an increase of on average +22.5% in systolic blood pressure, +21.0% in diastolic blood pressure and +32.6% in heart rate within 5 minutes after the stress exposure (Fig. 1I-K, condition x time *p* < 0.0001). There was no significant sex difference in the magnitude of stress-induced measures of affect or cardiovascular parameters (Fig. S1A-B).

### 1.2. Acute psychological stress selectively induces neuroendocrine reactivity

As expected, when compared to the control condition, exposure to stress was associated with a significant increase in plasma cortisol, peaking +4.3% above baseline 10 minutes after the onset of stress (effect of condition x time *p*=0.005, Fig. 2A), and a significant increase in plasma norepinephrine, peaking at +3.4% above baseline at 5 minutes after the onset of stress (effect of condition x time *p*=0.025, Fig. 2B). There were no significant condition differences in epinephrine, serotonin, or dopamine response (condition x time *p*’s > 0.05) (Fig. 2C-E). Results were retained in analyses that adjusted for baseline values (Fig. S2). There was no significant sex difference in the magnitude of stress-induced neuroendocrine reactivity (Fig S1C).

**Figure 2.**
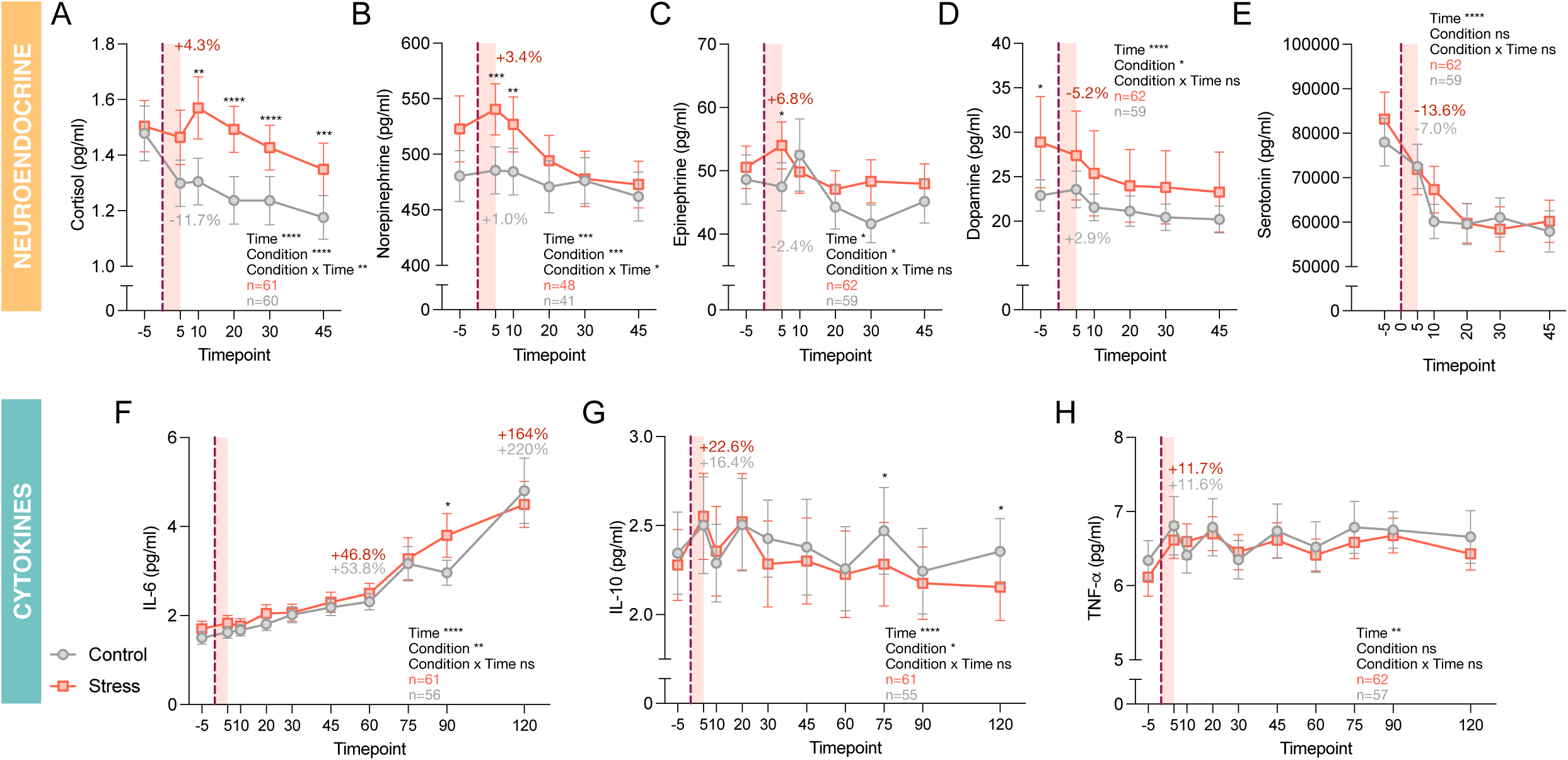
Neuroendocrine and immune activity in response to acute psychological stress. **(A-E)** Time courses of participants plasma cortisol and catecholamines levels in the stress and control conditions. Percentages represent change from baseline at **(A)** +10 min and **(B-E)** +5 min**. (F-H)** Time courses of participants plasma cytokine dynamics in the stress and control conditions. Percentages represent change from baseline at **(F)** +60 and +120 min, and **(G,H)** +5 min. Asterisks over symbols indicate a significant difference between control and stress values at the corresponding time point. (A-H) Data shown as average ± standard error of the mean (SEM). Effect sizes and p-values from mixed-effects analyses (of log-transformed values). *p<0.05, **p<0.01, ***p<0.001, ****p<0.0001.

### 1.3. Acute psychological stress does not lead to a selective increase in pro-inflammatory cytokines

Surprisingly, a gradual increase in circulating levels of IL-6 over time was detected in both stress and control conditions (time effect *p*<0.0001, Fig. 2F). There was also a significant main effect of time for trajectories of IL-10 and TNF-α (*p<*0.00001 and 0.002, respectively), although no clear pattern of response was observed across the task periods. When the trajectories were baseline-adjusted (Fig S2), a significant main effect of condition was observed on analysis of TNF-α with higher mean levels observed for the stress condition compared to the control condition. No significant interactions of time and condition were observed for any cytokine (condition x time *p*’s > 0.05, Fig. 2 F,G,H), nor did we observe significant sex difference in the magnitude of cytokine reactivity (Fig S1D).

### 1.4. Acute psychological stress does not lead to a selective increase in plasma or serum cf-mtDNA

Because cf-mtDNA content and biological interpretation depends on the biofluid studied^18,22,23^, we evaluated cf-mtDNA response to stress in both plasma and serum samples (Fig. 3A). Serum cf-mtDNA did not change significantly over time or in response to stress exposure (Fig. 3B). There was a main effect of time for plasma cf-mtDNA trajectories (*p*<0.00001) across both stress and control conditions, peaking at +22% above baseline at 10 minutes during stress visits (g=0.074, p=0.683, Fig. 3C) and +24% above baseline at 5 minutes during control visits (g=0.074, p=0.690). Under both conditions, cf-mtDNA levels decreased between the end of the stress task and 45 minutes, before rising to a local maximum at 75 minutes (stress: +22% above baseline, g=0.588, p=0.002; control: 8.5% below baseline, g=0.409, p=0.029), then decreasing gradually until the end of the time course at 120 minutes. No interactive effect of time and condition was observed for either serum or plasma cf-mtDNA trajectories.

**Figure 3.**
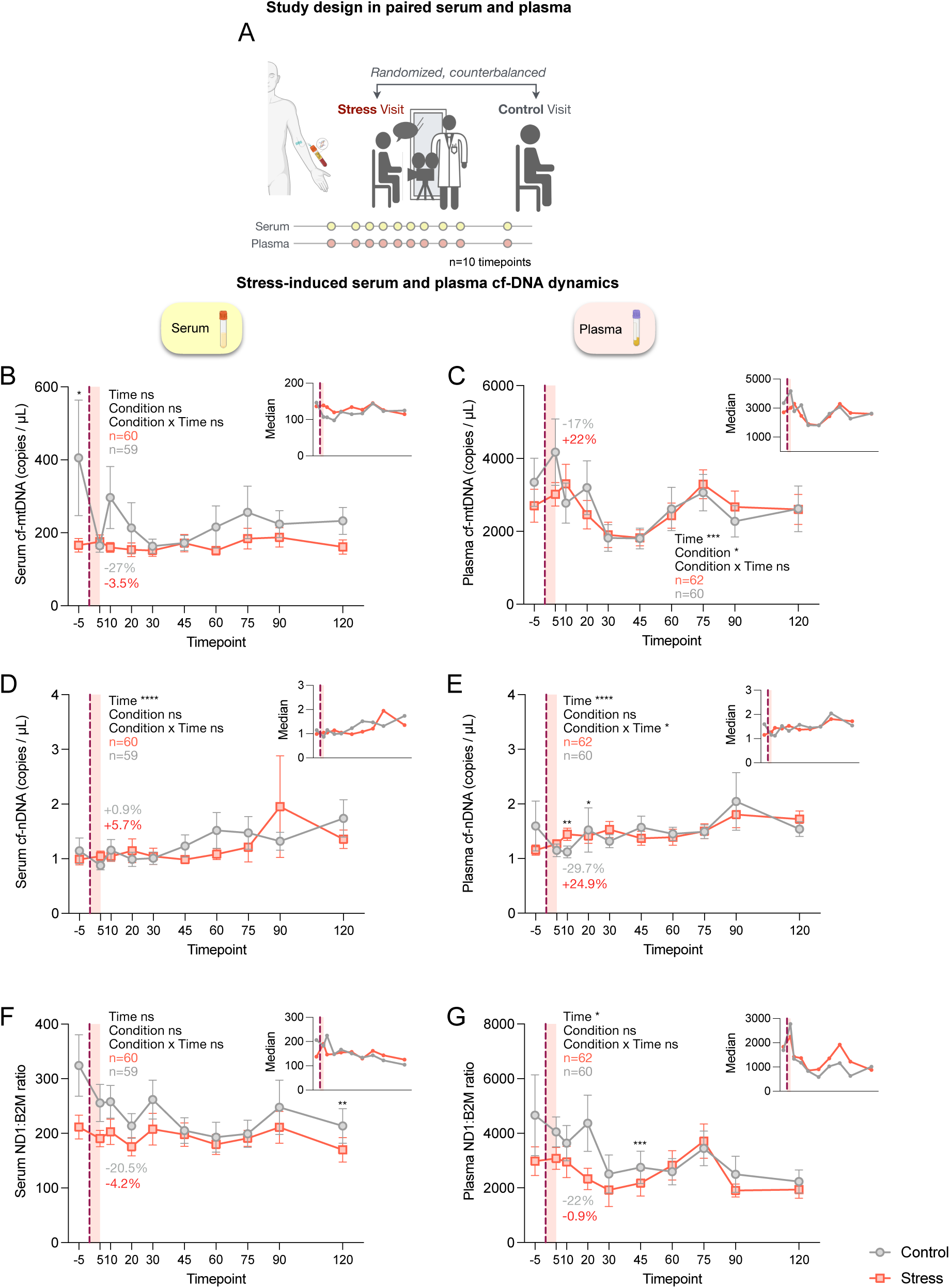
Cell-free DNA trajectories in plasma and serum in response to acute psychological stress. **(A)** During the stress and control visits, plasma and serum were collected simultaneously at 10 time points. Time courses of serum and plasma cf-DNA dynamics in the stress and control visits: **(B,C)** cf-mtDNA, **(D,E)** cf-nDNA, and **(F,G)** cf-mtDNA : cf-nDNA ratio (ND1:B2M). Values represent averages of all measurements in control and stress samples from 1st and 2nd visits. Percent changes between ‘-5 min’ and ‘+10 min’ are shown. Median values for each condition are shown in insets. Asterisks over symbols indicate a significant difference between control and stress values at the corresponding time point. **(B-G)** Data shown as average ± standard error of the mean (SEM). Effect sizes and p-values from mixed-effects analyses (of log-transformed values). *p<0.05, **p<0.01, ***p<0.001, ****p<0.0001.

To ensure the robustness of our plasma cf-mtDNA findings, we repeated the analysis using a different cf-mtDNA detection method and found strong agreement between the measures and similar cf-mtDNA trajectories (detailed results are shown in File S1. and Fig. S3). Similar results were found when values were adjusted for baseline levels (Fig. S4). We found a significant effect of time on baseline-corrected plasma cf-mtDNA values (p<10^-8^; Fig. S4D), but not baseline-corrected serum cf-mtDNA values (Fig. S4A), and no interactive effect of time and condition was observed for either sample type.

We also examined levels of cell free nuclear DNA (cf-nDNA). There was a significant main effect of time for both serum and plasma cf-nDNA levels (both *p* < 0.0001), with a gradual increase over time, by approximately 45% in serum and 18% in plasma on average (Fig. 3D,E). When cf-mtDNA (ND1) measurements were normalized to cf-nDNA (B2M) measurements to account for their potential shared origin and to quantify selective mitochondrial genome release, the results were similar to those of plasma cf-mtDNA. cf-mtDNA/cf-nDNA in serum did not change significantly over time, while time had a significant effect on ND1:B2M in plasma (*p* = 0.038) (Fig. 3F,G). No significant association between cf-mtDNA and cf-nDNA values was found in either serum or plasma (Spearman ρ ≤ 0.18; Fig. S5I,J).

No significant sex difference was found for the magnitude of cf-mtDNA reactivity to stress (Fig. S1E). However, when collapsed across conditions, men had higher *serum* cf-mtDNA levels than women (*p*=0.0002), while women had higher *plasma* cf-mtDNA levels than men (*p*=0.01) (Fig. S5A-B), again pointing to the distinct regulation of serum and plasma cf-mtDNA ^18,22,23^.

### 1.5. Plasma cf-mtDNA reactivity is not associated with autonomic reactivity

Next, we explored whether the cf-mtDNA trajectories to stress differed between individuals showing low or high autonomic responses to the stress task. For this purpose, we computed a composite autonomic response score (CARS) by averaging z-score transformed reactivities of heart rate, systolic and diastolic blood pressures, and norepinephrine, reflecting sympathetic nervous system activation. Results were median-split to classify participants as low or high autonomic responders (see Fig. 4A for the individual items). Serum or plasma cf-mtDNA trajectories did not significantly differ between high and low CARS groups (Fig. 4B-E, the plasma and serum cf-mtDNA trajectories by stress physiological and mood reactivity are shown in Fig. S2 and Fig. S6-8) suggesting that sympathetic nervous system stress reactivity might not contribute to variation in blood cf-mtDNA.

**Figure 4.**
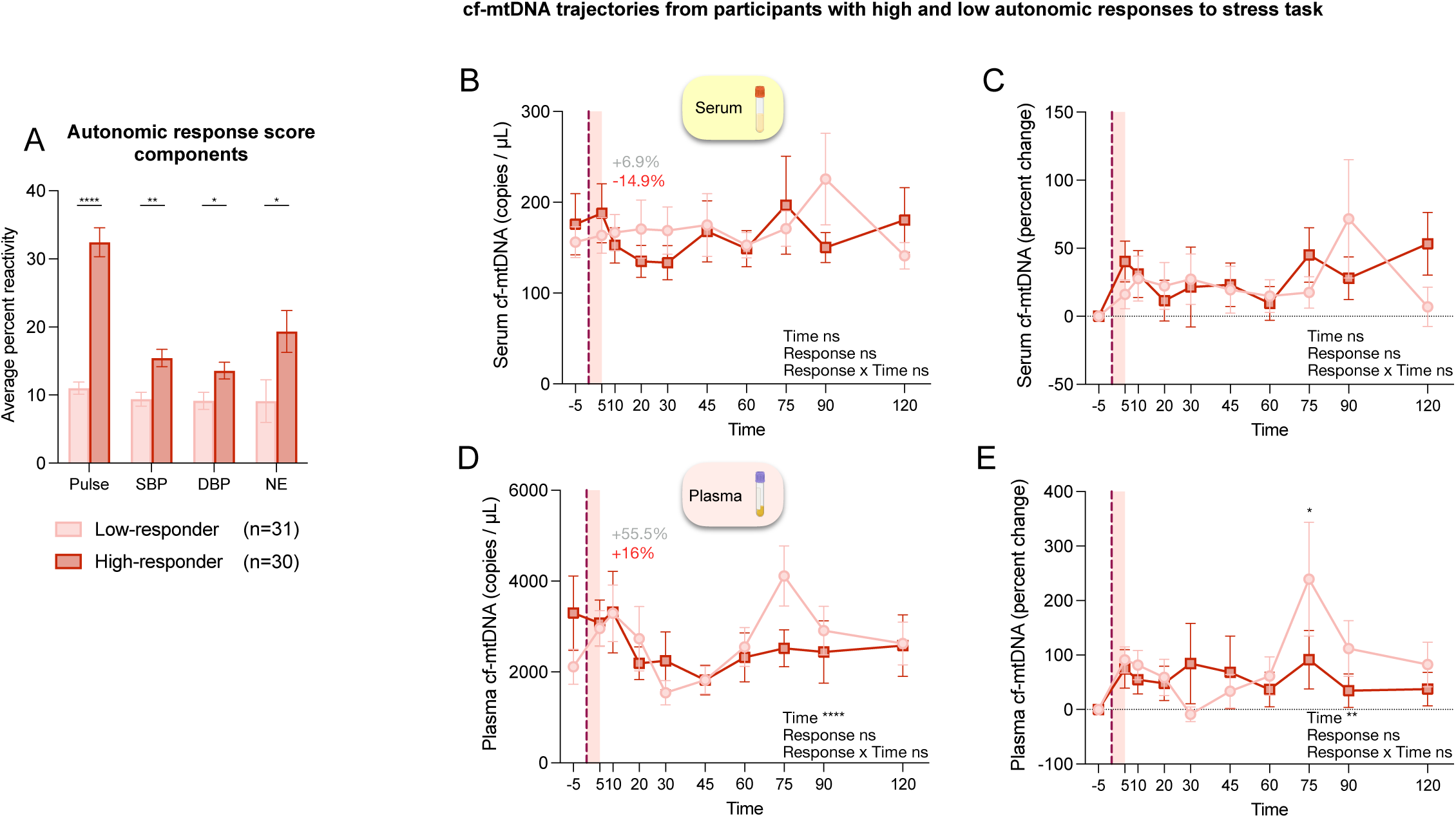
cf-mtDNA trajectories in response to stress by participants autonomic responses. To evaluate participants autonomic response to the stress task, we created a composite autonomic response score (CARS). For each participant, the CARS was calculated by averaging the z-score transformed reactivities of pulse rate, systolic blood pressure (SBP), diastolic blood pressure (DBP), and norepinephrine (NE) during the stress visit. Time windows used for calculating variable reactivities are defined in Supplemental Table 2. **(A)** Bar plot illustrating the individual component of the CARS for participants whose CARS were above and below the median CARS value (low-responders and high-responders, respectively). Time course of serum cf-mtDNA during the ‘stress’ visits for low and high autonomic responders shown as **(B)** copies of ND1 / µL and **(C)** percent change from baseline**. (D,E)** Same as in A,B for plasma cf-mtDNA. Percentages shown in **(B,D)** represent change in average cf-mtDNA abundance (copies / µL) between +10 and -5 min. Asterisks over symbols indicate a significant difference between low and high responder values at the corresponding time point. Data shown as average ± standard error of the mean (SEM). Effect sizes and p-values from **(A)** unpaired t-tests and Holm-Šidák’s multiple comparisons tests and **(B-E)** mixed-effects analyses (of log-transformed values). *p<0.05, **p<0.01, ***p<0.001, ****p<0.0001.

### 1.6. Plasma cf-mtDNA reactivity is not associated with pro-inflammatory cytokines reactivity

Finally, we examined whether the magnitude of reactivity in response to the stress condition was correlated between plasma cf-mtDNA and pro-inflammatory cytokines. Contrary to our original hypothesis, no significant association was found between stress-induced changes in plasma cf-mtDNA, IL-6, IL-10 and TNF-α (Fig S4K-M), suggesting that cf-mtDNA is not associated with concomitant change in circulating inflammatory mediators.

## 2. Discussion

In this crossover experimental trial, we investigated blood cf-mtDNA dynamics in response to a laboratory psychosocial stress condition and examined whether responses relate to mood, cardiovascular, neuroendocrine and pro-inflammatory cytokines reactivity. Our findings show that while the socio-evaluative stress condition induced the expected mood, cardiovascular, and neuroendocrine responses, validating the task as an effective stressor, acute mental stress did not produce the expected pro-inflammatory cytokines or cf-mtDNA responses above the control condition.

The current findings confirm previously observed increases in IL-6 following exposure to a socio-evaluative stressor. Indeed, results of a meta-analysis of IL-6 responses to acute laboratory stressors support moderate to large increases in circulating levels at 90 and 120 minutes following stress ^1^. However, contrary to expectations, the current results showed this pattern of response following both the stress and the control condition. To date, few studies examining cytokine responses to acute stress have included a control condition ^1^, some ^24,25^ have found a selective effect of psychological stress on circulating pro-inflammatory cytokines increase. The current results raise the possibility that previously observed increases in IL-6 following acute laboratory challenge do not result from stress exposure *per se* but from factors common across both conditions, such as diurnal variation in cytokine concentrations ^26^, a local response to the presence of an intravenous catheter ^27^, prolonged sitting ^28,29^, the arousal of attending a lengthy study session, separation from cell phone, boredom (we saw a decrease in well-being in both the control and stress conditions) or other non-specific aspects of the protocol. Further research is needed to better understand the reasons for increased circulating levels of IL-6 in the control condition, especially given that IL-6 is widely recognized to increase in response to acute psychological stress.

In our prior uncontrolled study, we examined serum cf-mtDNA levels before and 30 minutes after the same 5-minute socio-evaluative stressor employed in the current study. These initial results showed a 1-2-fold increase in serum cf-mtDNA 30 minutes post-stress (n=50) across two sessions separated by one month ^9^. Contrary to expectations, we did not replicate these findings in the current study (no effect of time was found) or see any clear difference in the pattern of reactivity between the stress and control conditions. The reasons for inconsistent findings across our two studies remain unclear but could be influenced by methodological (e.g. samples were stored in the freezer for a longer period in the first study) or participant-related differences (the population was older in our previous study). Given evidence that levels of cf-mtDNA differ between serum and plasma ^18^, we extended our prior work to also examine plasma levels of cf-mtDNA in the current study. In plasma, cf-mtDNA exhibited significant temporal variation in both conditions. In the stress condition, plasma cf-mtDNA peaked by 22-24% at 5-10 minutes post-stress, decreased until 45 minutes, rose again at 75 minutes, and then gradually declined until 120 minutes. These results align with prior findings showing a 60% increase in plasma cf-mtDNA levels 15 minutes post-socio-evaluative stress in an uncontrolled study of healthy men (n=20) ^8^. A more recent study by the same group reported a significant 70% increase in plasma cf-mtDNA levels 45 minutes post-stress (n=42) ^10^. However, since we observed no significant differences in the variation in plasma cf-mtDNA between the stress and control conditions, our results suggest that the observed cf-mtDNA increase might not be attributable to psychological stress exposure. As for the increased in levels of IL-6 discussed above, this pattern of results suggests that other factors associated with the laboratory visit, such as catheter placement, might contribute to the cf-mtDNA elevation observed in this study.

An aim of the current study was to examine factors that relate to and may possibly be involved in stress-related changes in circulating levels of cf-mtDNA. Although we did not observe clear stress effects, for the plasma measure of cf-mtDNA reactivity, we did observe a main effect of time across both conditions. Thus, we examined associations of cardiovascular, neuroendocrine and psychological factors with this temporal response. Although prior work from our group showed that serum cf-mtDNA responses to acute socio-evaluative stress were influenced by baseline mood and cardiovascular state, ^16^ we did not observe an association between plasma cf-mtDNA reactivity and markers of autonomic activity or mood status in response to the tasks. Our earlier findings linked larger stress-related serum cf-mtDNA responses to higher baseline heart rates, smaller decreases in heart rate variability, and larger increases in fatigue ^16^. The inconsistencies between our two studies may be attributable to the distinct biological properties of serum and plasma cf-mtDNA. Future studies with larger sample sizes should investigate the factors driving plasma cf-mtDNA variation to help understand its potential physiological significance.

Finally, we investigated the association between the magnitude of pro-inflammatory cytokine and plasma cf-mtDNA responses across the two laboratory sessions. Prior studies suggest that cf-mtDNA might be pro-inflammatory. Injection of purified mtDNA into mice or its exposure to cultured cells can trigger the production of proinflammatory cytokines ^12,14^. In humans, a study found that older adults with increased circulating cf-mtDNA exhibited higher levels of the proinflammatory cytokines IL-6 and TNF-α ^13^. Additionally, in isolated monocytes, mtDNA acts as an adjuvant, enhancing TNF-α production ^13^. In the present study, we found no evidence of a significant association between the pattern of cf-mtDNA and pro-inflammatory cytokines responses over time. For cf-mtDNA to elicit a pro-inflammatory response, it must be accessible to cellular receptors ^30-32^. However, in healthy individuals, most cf-mtDNA (>90%) is encapsulated within vesicles or whole mitochondria ^31,33^, which likely limits its inflammatory potential. This suggests that cf-mtDNA might serve non-inflammatory roles, such as facilitating intercellular transfer of mitochondria or mtDNA to modulate recipient cells’ bioenergetic capacity. While this remains to be confirmed by future studies, cf-mtDNA response may represent a healthy physiological mechanism rather than a pro-inflammatory reaction, highlighting the potential for cf-mtDNA to fulfill roles beyond inflammation, including cellular communication and adaptation.

Although our study was strengthened by the inclusion of a control condition, it has several limitations. The sample size was too small to adequately assess predictors of inter-individual differences in the magnitude of cf-mtDNA responses to stress. Larger studies are needed to understand why some individuals exhibit large cf-mtDNA responses while others show none or even negative cf-mtNDA reactivity. Additionally, the external validity is limited as our sample consisted solely of healthy individuals ages 20–50. One key technical challenge is the significant diversity and heterogeneity of mitochondria-like particles in plasma and serum ^34^. Ideally, future studies should isolate, purify, and analyze specific microparticles (including whole mitochondria ^33^, platelet-derived fragments, and exosomes) for mtDNA content. However, technical constraints and the study’s scope prevented such detailed analyses. Future research would benefit from greater analytical specificity to improve the precision and interpretation of cf-mtDNA measurements.

### Conclusions

Our study contributed to the growing understanding of pro-inflammatory cytokines and cf-mtDNA dynamics in response to acute psychosocial stress. While we observed significant temporal variation in plasma pro-inflammatory cytokines (IL-6, IL-10, and TNF-_J) and plasma cf-mtDNA over a 2-hour period of sitting with an intravenous catheter, the lack of differences in these responses between stress and control conditions suggests that factors beyond psychological stress, such as procedural elements of the laboratory setting, may drive these changes. The distinct behaviors of cf-mtDNA in serum and plasma underscore the need for biofluid-specific analyses in future research, and for studies in other biofluids like saliva ^35^. Additionally, the absence of an association between cf-mtDNA and pro-inflammatory cytokines suggests a non-inflammatory role for cf-mtDNA ^36^, potentially related to intercellular communication and bioenergetic regulation. Future studies with larger, more diverse samples and refined methodological approaches will be essential to further elucidate the physiological and long-term health significance of cf-mtDNA reactivity in humans.

## 3. Material and methods

### 3.1. Participants

Healthy adults were recruited though advertisements in the community and via a University of Pittsburgh participant registry to participate in The Mitochondria and Psychological Stress (MaPS) study (clinicaltrials.gov #NCT04078035). All enrolled participants provided written informed consent as approved by the Institutional Review Board of the University of Pittsburgh. Eligibility criteria were (1) 20-50 years of age; (2) for females, regular menstrual cycles over the past year, not pregnant or lactating; (3) generally physically healthy (no reported history of chronic immune, metabolic, nervous system, endocrine, or mitochondrial diseases); (4) not taking medications targeting the central or peripheral nervous, or neuroendocrine systems; (5) no reported history of psychotic illness or mood disorder; (6) resting blood pressure<140/90 mmHg; (7) not a current smoker or illicit drugs user; (8) weight>110lbs and BMI<30; and (9) fluent in English (used everyday in speaking and reading for at least 10 years). Recruitment occurred between July 2020-December 2022. 124 individuals were screened for participation among which 88 were eligible and came for a consent visit. At the consent visit, 4 individuals were excluded for health conditions (high blood pressure or high BMI). Participants were rescheduled if they reported (1) having an infection, cold or flu or were taking antibiotics or anti-inflammatories, or having received a vaccination or tattoo in the two weeks before the visit; (2) presented with symptoms of an infection on the day of the visit; or (3) reported taking glucocorticoids in the 24 hours preceding the visit. For female participants, both visits were scheduled during the luteal phase (21-28 days post menstruation) of their menstrual cycle to control for hormonal vairations. A total of 84 participants were enrolled, of those 12 did not complete the first visit because the IV insertion failed. Ultimately, 72 participants completed the first visit. Of those, 69 returned for the second visit (n=3 lost at follow-up), the IV insertion failed on 17, 52 participants completed visit 2.

### 3.2. Stress Reactivity Protocol

Participants attended up to two laboratory sessions spaced at least one month apart at the Behavioral Physiology Laboratory, University of Pittsburgh. Both sessions started in the afternoon (between 1:00-3:00 PM) to avoid the confound of the cortisol awakening response ^37^ and accurately capture the stress-induced cortisol response. Participants were asked to avoid alcohol and vigorous exercise for 24 hours, non-prescription drugs for two days, and food and caffeine for 3 hours prior to each session. During the “stress” session, participants completed a 5-minute socio-evaluative speech task as described previously ^9,21^. During the “control” session, participants were not exposed to the stressor and rested quietly for the 5-minute task period. The order of sessions was randomized and counterbalanced across participants. On arrival at the first visit, the research nurse conducted a medical history and medication use screen, and recorded participants’ resting blood pressure, height, weight, and percent body fat. Participants who were confirmed to be eligible were then asked to complete demographic and psychosocial questionnaires. At both visits, participants were seated and instrumented for the psychophysiologic testing session. This included insertion of an intravenous catheter into the antecubital fossa of one arm for the collection of blood samples and placement of a cuff on the opposite arm for the automated measurement of blood pressure and heart rate (Dinamap). Participants then sat quietly for a 30-minute baseline (habituation) period before completing either the stress task or the control rest period. After the task, participants watched a neutral wildlife documentary for a 120-minute recovery period. Blood samples were drawn 5 minutes before the start of the task and 5, 10, 20, 30, 45, 60, 75, 90, and 120 minutes after the beginning of the task. Blood pressure and heart rate were measured every 2 minutes during the last 10 minutes of the baseline period and twice at the same time as each blood draw thereafter. Except when performing the speech task, participants were left alone as much as possible. On occasion, the study nurse had to use a tourniquet (33% of total visits) or re-insert or reposition the catheter (8% of total visits).

### 3.3. Questionnaires and anthropometric measures

Participants completed a 38 question Profile of Mood States (POMS) ^38,39^ questionnaire 5 minutes before the start of the task and 5, 10, and 20 minutes after the beginning of the task. The responses to these questions were converted to seven POMS scores (fatigue, anger, anxiety, depression, vigor, wellbeing, and calmness).

### 3.4. Blood collection and processing

Serum and EDTA plasma were collected in vacutainers (BD cat. #367812 and #366643) at every timepoint (10). Serum was collected before plasma at each time point. After collection, plasma was immediately centrifuged at 1,000 x *g* for 5 minutes at room temp and then temporarily stored at 4° C, while serum samples were incubated at room temperature for 30 minutes to allow clotting prior to centrifugation and refrigeration. Supernatants (80%) from plasma and serum tubes were transferred to conical tubes and centrifuged a second time at 2,000 x *g* for 10 minutes at 4° C. Supernatants (90%) from conical tubes were transferred to new conical tubes and homogenized, then aliquoted and frozen at -80° C.

### 3.5. Cortisol and catecholamines assay

Plasma cortisol and catecholamine levels were assessed in duplicate from blood samples drawn at -5, 5, 10, 20, 30, and 45 minutes relative to the start of the task period. Cortisol was assessed using a cortisol ELISA kit (R&D Systems, cat. #KGE008B) run according to manufacturer’s directions. Intra- and inter-assay CVs were 5.68% and 5.54%, respectively. Measurements of norepinephrine, epinephrine, dopamine, and serotonin by liquid chromatography tandem mass spectrometry (LC-MS/MS) were carried out by the Biomarkers Core Laboratory at the Irving Institute for Clinical and Translational Research (Columbia University). Samples were spiked with deuterated internal standards and analytes were extracted using protein precipitation and derivatization with dansyl chloride. Analytes were further processed using ethyl acetate liquid-liquid extraction and suspended in acetonitrile prior to MS. Chromatography was run on a Waters ACQUITY UPLC HHS C18 column (2.1 x 100 mm, 1.8 µm) at 40 °C. Gradient elution was performed with water and acetonitrile with 0.1% formic acid mobile phases at a flow rate of 300 µL / minute. LC-MS/MS analysis with positive ESI and multiple reaction monitoring (transitions: norepinephrine 869.2>170.3; epinephrine 883.3>170.2; dopamine 853.30>170.37; serotonin 643.29>146.05) was performed on a Waters Xevo TQS MS (Waters, Milford MA, USA).

### 3.6. Pro-inflammatory cytokines assay

Pro-inflammatory cytokines (IL-6, IL-10, and TNF-_J) were measured using the Ella automated immunoassay platform (ProteinSimple, Biotechne) as described previously ^40^. All available time points for each participant were assayed on the same plate. Intra- and inter-assay CVs ranged from 2.64-6.6% and 4.1-10.0%, respectively.

### 3.7. cf-DNA quantification

Cell-free nuclear and mitochondrial DNA in serum and plasma were quantified using methods that have been described in detail (Ware et al., 2020). Briefly, 150 µL of plasma supernatant or 75 µL of serum supernatant was combined with 5.7 µL of 20 mg/mL Proteinase K and 7.5 µL of 20% SDS on a deep-well plate, which was incubated at 70° C for 16 hours. After cooling to room temp, plates were centrifuged for 1 minute at 500 x *g*. MagMAX cell-free DNA lysis/binding solution (125 µL for serum or 250 µL for plasma; ThermoFisher cat. #A33600) and magnetic beads (5 µL for serum or 7.5 µL for plasma; Dynabeads MyOne Silane; ThermoFisher cat. #37002D) were added to each sample. Nucleic acids in samples were washed three times using a KingFisher Presto magnetic particle processor (ThermoFisher cat. #5400830; Wash 1: 265 µL of MagMAX cell-free DNA wash solution / well for serum or 375 µL for plasma, ThermoFisher cat. #A33601; Wash 2: 475 µL 80% ethanol / well for both sample types; Wash 3: 200 µL 80% ethanol / well for both sample types) and finally eluted into MagMAX cell-free DNA elution solution (60 µL for both sample types; ThermoFisher cat. #A33602) on a 96-well microplate. Isolated nucleic acids were split into two 96-well plates and stored at -20° C. After thawing, TaqMan-based qPCR reactions were prepared in triplicate on 384-well plates with 3.2 µL of template DNA, 4 µL of 2x Luna universal qPCR master mix (New England Biolabs cat. #M3003), and 0.4 µL of 20x primer / probe solutions for mitochondrial target gene ND1 and nuclear target gene B2M. qPCR plates were also loaded with a seven-step four-fold serial dilution of human placenta DNA that had been quantified previously using digital droplet qPCR (ddPCR). qPCR was run on a QuantStudio 5 real-time PCR system. Means of triplicate cycle threshold values (C_T_) were compared to measurements of the standard curve to quantify copies of target genes per µL of sample.

The field of cf-mtDNA detection is still in development, and to ensure the robustness of our results, we used an additional method to measure cell-free DNA in plasma samples, analyzed by a different lab. For this purpose, we used the ‘MitoQuicLy’ method (Michelson et al., 2023) with several modifications. Briefly, 10 µL of each plasma supernatant was combined with 190 µL of lysis buffer (114 mM Tris-HCl, pH 8.5; 6% Tween-20; 200 µg / mL Proteinase K) on two replicate 96-well plates, which were heated on a thermocycler to 55° C for 16 hours and 95° C for 10 minutes. Lysate plates were also loaded with four reference plasma samples. Samples from the same participant were processed and analyzed together to prevent batch effects from influencing within-participant comparisons. Lysates were either held at 4° C until qPCR analysis within few hours after lysis, or frozen and stored at -80° C until analysis. 8 µL of each replicate lysate was combined with 12 µL of qPCR buffer (TaqMan Universal MasterMix Fast; 10 µM of forward and reverse qPCR primers for mitochondrial target gene ND1 and nuclear target gene B2M; 5 µM of qPCR probe for each target gene) in triplicate on a 384-well plate. Each qPCR plate was also loaded, in triplicate, with an eight-step 1:4 serial dilution of purified fibroblast DNA that had been quantified previously using ddPCR. qPCR was run on a QuantStudio 7 Flex real-time PCR system. The medians of triplicate C_T_ measurements of each lysate were compared to the median C_T_s of each dilution of the standard curve from corresponding qPCR plates to estimate copies of target genes per µL. To correct for plate effects, a correction factor was calculated for each pair of replicate plates by dividing the mean of its four reference plasma measurements by the experiment-wide mean of reference plasma measurements. Sample measurements were divided by their plate’s correction factor to determine adjusted values. The means of each sample’s adjusted replicate values are presented in figures and were used in statistical analyses. Plates were re-analyzed, starting from the lysis stage, if the median coefficient of variance (CV) between replicate measurements was greater than 4.0%. Five plates of samples were re-analyzed based on this condition. Sequences of primers and probes used in qPCR are available in Supplemental Table S1.

### 3.8. Data processing

For measurements recorded at multiple time points during stress and control visits, single reactivity values were calculated as follows: POMS mood reactivities (anger, anxiety, depression, vigor, wellbeing, and calmness) were defined as the difference between baseline and the value reported immediately after the stress task (5 minutes from start). POMS fatigue reactivity was calculated as the absolute value of the difference between baseline and +5 minutes. Cardiovascular (heart rate, systolic blood pressure, diastolic blood pressure), neuroendocrine (cortisol, norepinephrine, epinephrine, serotonin, dopamine), immune (IL-6, IL-10, TNF-α) and cf-mtDNA reactivity was defined as the percent change from baseline to the maximum value observed within a specific time window post-task. Baseline was defined as the - 5 minute time point for all variables except for cardiovascular variables, for which the average of five measurements recorded every 2 minutes during the last 10 minutes of the baseline period was used. Cardiovascular measurements at subsequent time points represent the average of two values recorded close together. The windows for calculating percent reactivity were defined as follows: cardiovascular measures: 0-20 minutes following start of task; norepinephrine and epinephrine: 5-10 minutes; cortisol and dopamine: 5-30 minutes; proinflammatory cytokines and cf-mtDNA: 5-120 minutes (See Supplemental Table S2 for more details).

We calculated a composite autonomic response score (CARS), by averaging the z-score transformed stress reactivities of pulse rate, diastolic blood pressure, systolic blood pressure, and norepinephrine. For this composite score, and for each individual physiological variable and POMS mood score, participants with z-score transformed stress reactivities above or below median were designated as ‘low-responders’ or ‘high-responders’, respectively.

### 3.9. Statistical analysis

To compare trajectories of variables measured at multiple time points between conditions (i.e., data from stress versus control visits, or participants who had high or low autonomic responses during stress visits) linear mixed-effects models were fit to the data using the “lmerTest” package in R (version 4.3.0), with subject as random effect and condition and time point as fixed effects. cf-mtDNA, cf-nDNA, all cytokines and neuroendocrine variables were log transformed prior to mixed-effects modeling. The following analyses were run using GraphPad Prism version 10.4.0 (GraphPad Software, Boston, Massachusetts USA): Computation of the relative likelihood of data having a normal or lognormal distribution ^41^, Spearman’s rank correlation to evaluate associations between continuous variables, and unpaired t-tests and Mann-Whitney tests to compare values between groups with and without normally distributed values, respectively.

## Acknowledgements

The MaPS study was supported by the NIH grant R01MH119336 to M.P., A.M., and B.K.

## Data availability

Data will be made available upon request.

## Authors contributions

A.L.M., M.P., B.A.K. and C.T. designed the study. A.L.M. supervised data collection and K.S. recruited participants and collected study visits data and samples. D.S. and A.W. performed the cf-mtDNA assays. A.L.M., S.L. and C.T. designed the data analysis plan. C.T. and D.S. ran the statistical analysis and prepared the manuscript. All authors reviewed the final version of the manuscript.

## Supplements

**Supplemental Figure 1.**
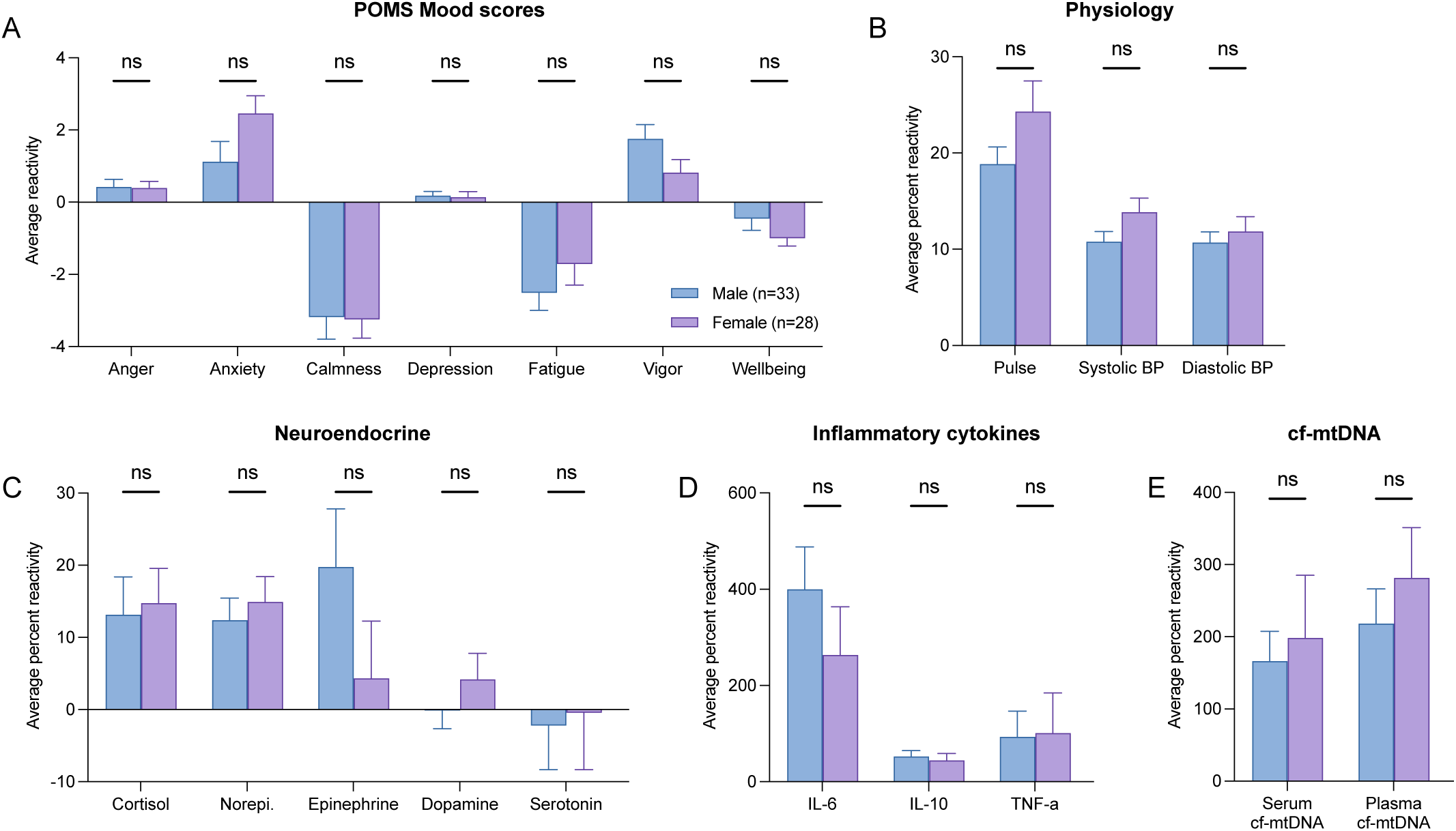
Comparison of responses to stress task by sex. **(A-E)** Bar graphs representing average **(A)** mood, **(B)** physiological, **(C)** neuroendocrine, **(D)** cytokine, and (**E)** cf-mtDNA responses during stress visits in male and female participants. Data shown as average ± standard error of the mean (SEM). Effect sizes and p-values from unpaired t-tests. *p<0.05, **p<0.01, ***p<0.001, ****p<0.0001.

**Supplemental Figure 2.**
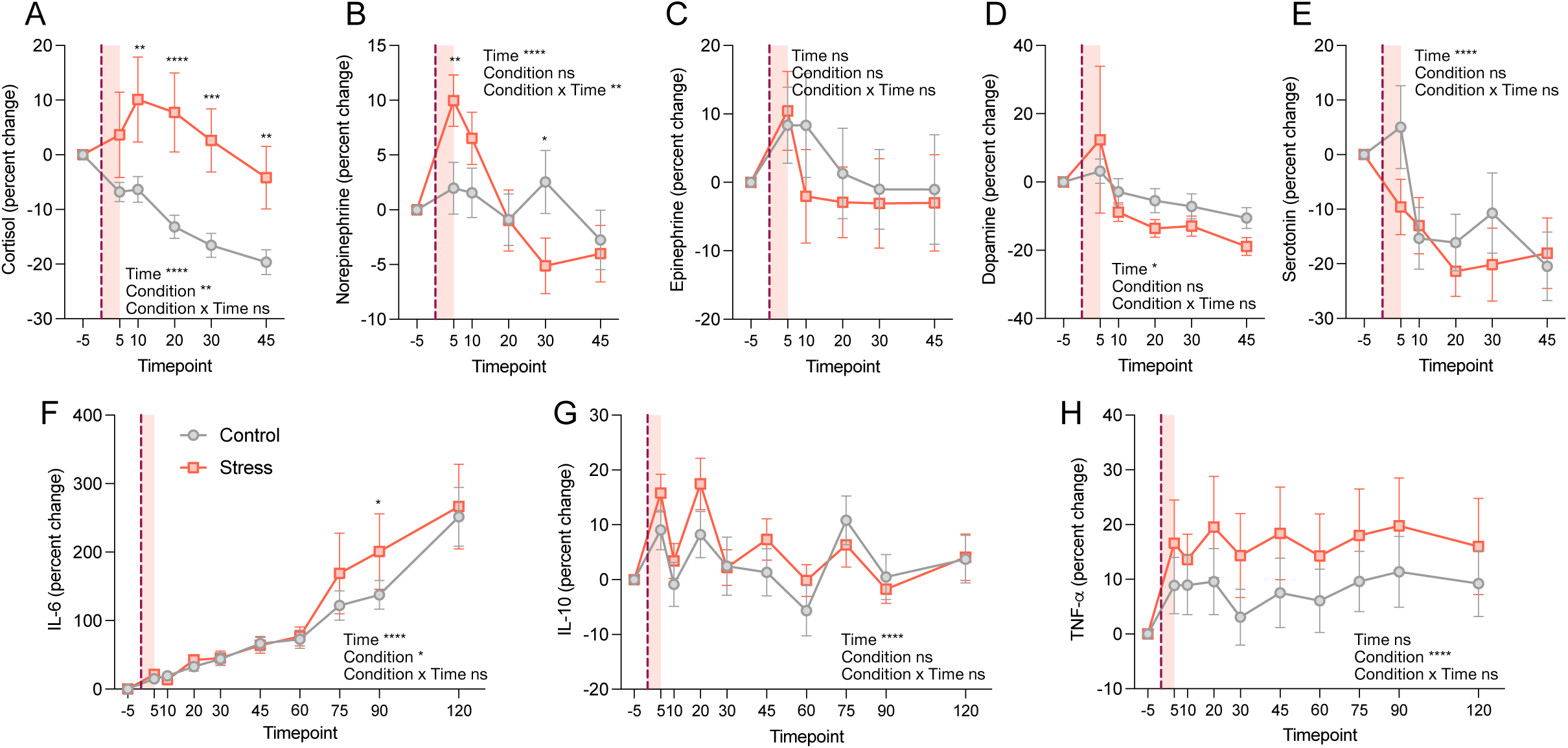
Neuroendocrine and immune stress reactivity shown as percent change from baseline. **(A-E)** Time courses of participants plasma cortisol and catecholamines levels in the stress and control conditions. Values represent the average percent change between baseline (-5 min) and subsequent time points. **(F-H)** Time courses of participants plasma cytokine dynamics in the stress and control conditions shown as percent change from baseline. Asterisks over symbols indicate a significant difference between control and stress values at the corresponding time point. Data shown as average ± standard error of the mean (SEM). Effect sizes and p-values from mixed-effects analyses. *p<0.05, **p<0.01, ***p<0.001, ****p<0.0001.

**Supplemental Figure 3.**
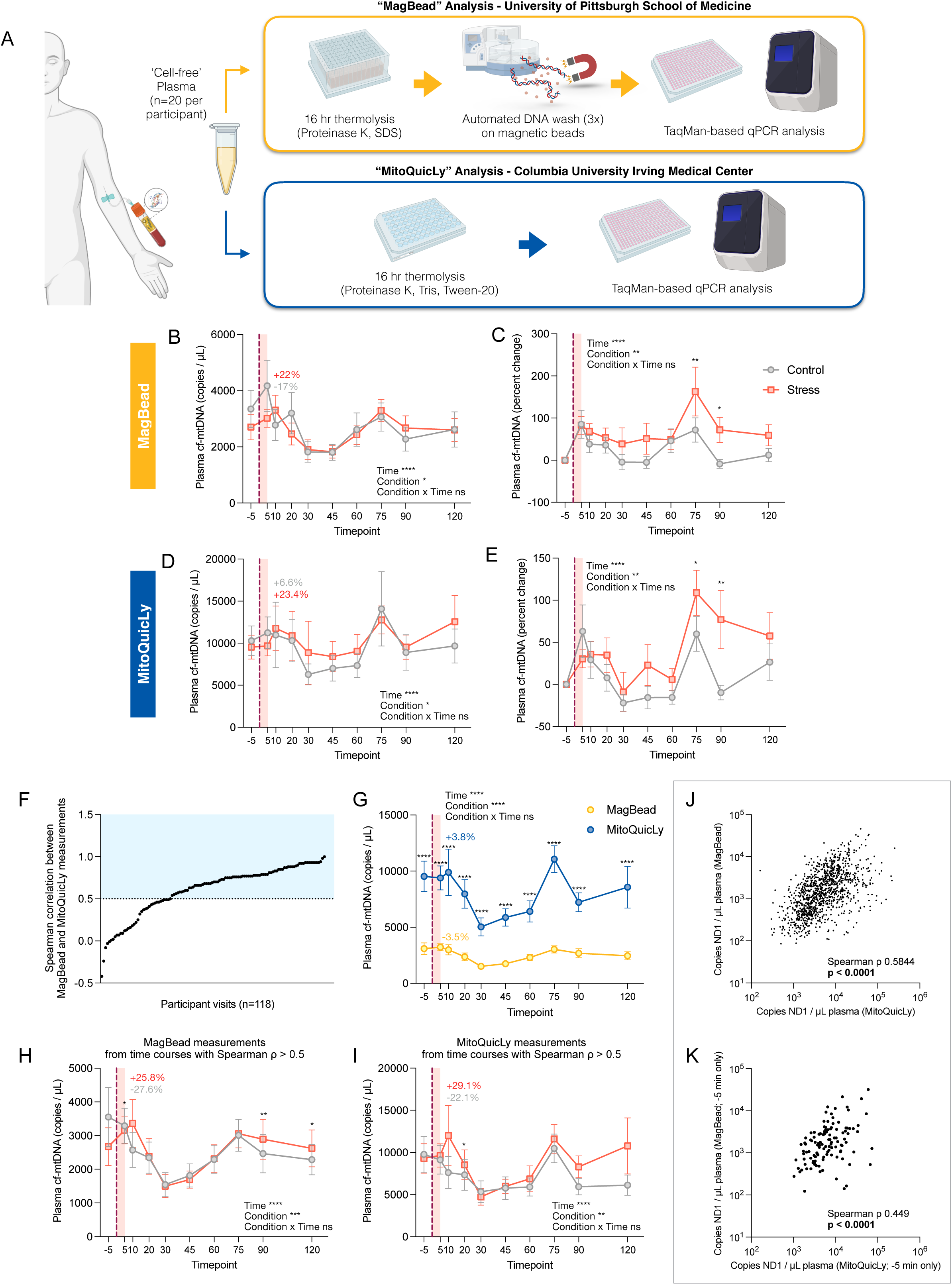
Comparison of plasma cf-mtDNA levels measured with the “MagBead” and “MitoQuicLy” methods. **(A)** Overview of “MagBead” and “MitoQuicLy” methods. Plasma cf-mtDNA was quantified using both approaches. Each method began with thermolysis for 16-hours, but with different lysis buffer formulations. In the MagBead approach, DNA in lysates was washed several times using an automatic magnetic bead-based system prior to qPCR analysis, while the MitoQuicLy approach had no intermediate step between lysis and qPCR. **(B-E)** Time courses of plasma cf-mtDNA levels derived from MagBead and MitoQuicLy methods shown as **(B,D)** copies / µL and **(C,E)** percent changes from values at ‘-5’ time point. **(F)** Distribution of the association (assessed with Spearman ρ) between MitoQuicLy and MagBead measurements from individual time courses. The blue-shaded area highlights time courses showing good agreement when assayed using the two methods (ρ ≥ 0.5). **(G)** Time courses of plasma cf-mtDNA values measured by the MagBead and MitoQuicLy methods. **(H,I)** Time courses of plasma cf-mtDNA levels showing good agreement (ρ ≥ 0.5, blue area in (F)) between the two methods for **(H)** MagBead or **(I)** MitoQuicLy. **(J,K)** Scatterplots of the association between plasma cf-mtDNA levels measured by the MagBead and MitoQuicLy approaches for **(J)** all timepoints and **(K)** baseline values. Percentages shown in (B-E,G-I) represent change in average cf-mtDNA abundance (copies / µL) between +10 and -5 min. Asterisks over symbols on trajectory plots indicate a significant difference between control and stress values at the corresponding time point. (B-E, G-I) Data shown as average ± standard error of the mean (SEM). Effect sizes and p-values from (B-E, G-I) mixed-effects analyses (of log-transformed values) and (J,K) Spearman rank correlation. *p<0.05, **p<0.01, ***p<0.001, ****p<0.0001.

**Supplemental Figure 4.**
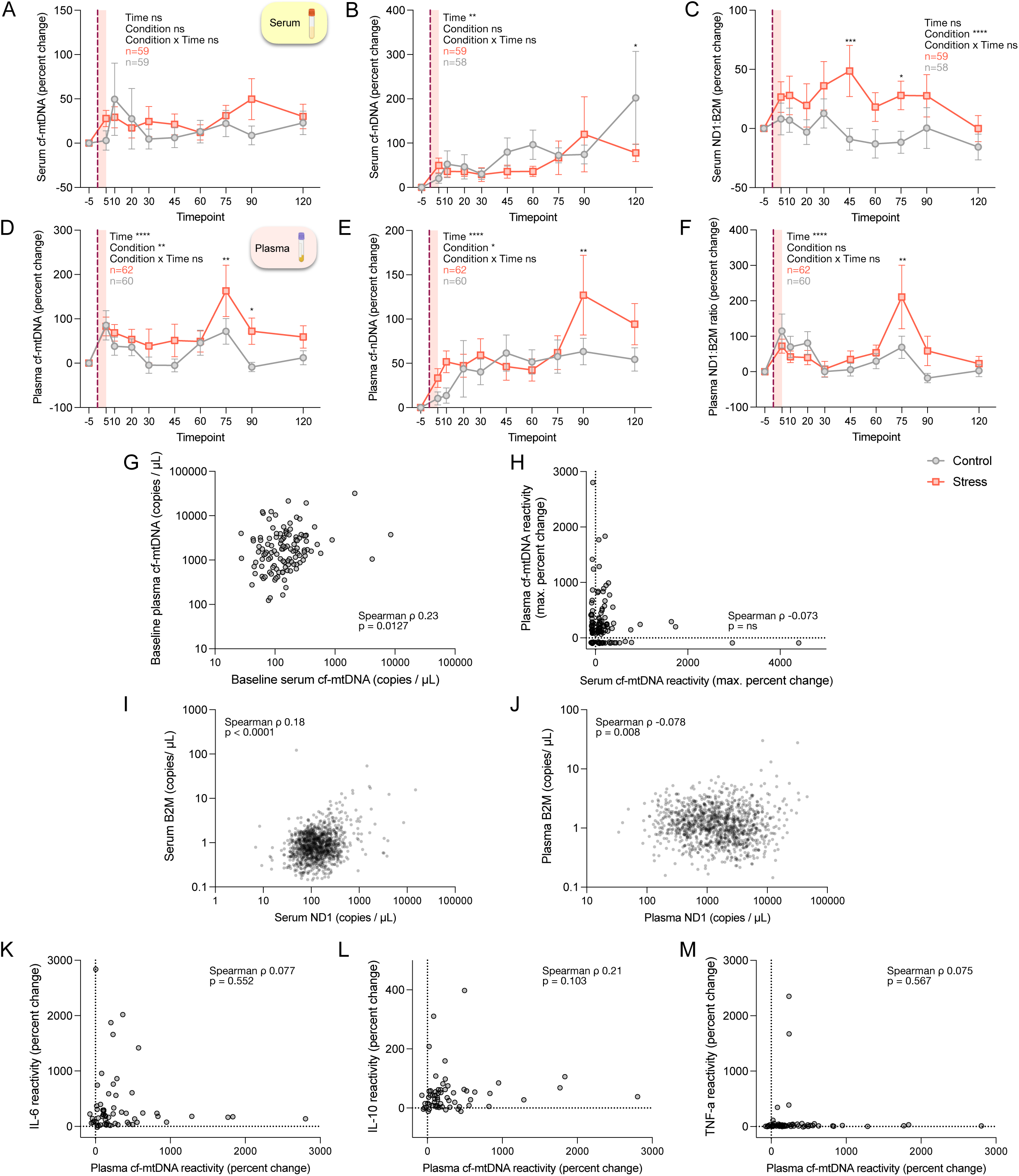
Baseline normalized plasma and serum cf-DNA trajectories and comparisons of cf-mtDNA levels between sample types. **(A-F)** Time courses of serum and plasma cf-DNA dynamics in the stress and control visits, where values represent the average percent change between -5 min and subsequent time points. Trajectories presented include baseline normalized **(A,D)** cf-mtDNA, **(B,E)** cf-nDNA, and **(C,F)** cf-mtDNA : cf-nDNA ratio (ND1:B2M)**. (G,H)** Scatterplot of the association between **(G)** baseline cf-mtDNA and **(H)** cf-mtDNA reactivity in plasma and serum. Cf-mtDNA reactivity was defined as the percent change between -5 min and the largest value measured over the remainder of the time course. **(I,J)** Scatter plots illustrating the relation between ND1 and B2M measured in each **(I)** serum and **(J)** plasma sample. **(K-M)** Scatter plots illustrating the relation between the reactivities of cf-mtDNA and **(K)** IL-6, **(L)** IL-10, and **(M)** TNF-ɑ during stress visits, where reactivity is defined as the percent difference between baseline and the largest value measured in subsequent time points. Asterisks over symbols indicate a significant difference between control and stress values at the corresponding time point. (A-F) Data shown as average ± standard error of the mean (SEM). Effect sizes and p-values from **(A-F)** mixed-effects analyses and **(G-M)** Spearman correlations. *p<0.05, **p<0.01, ***p<0.001, ****p<0.0001.

**Supplemental Figure 5.**
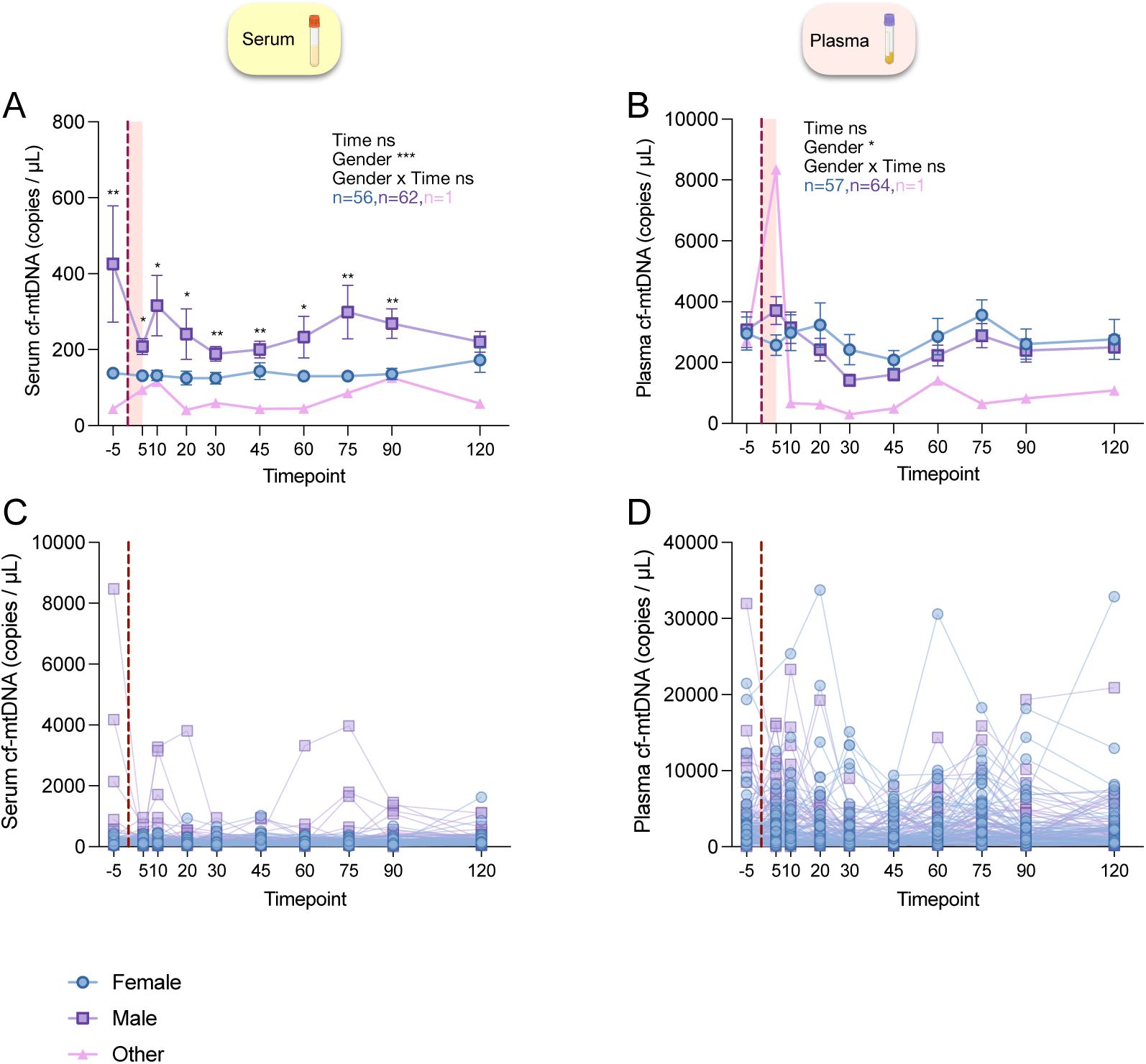
Cell-free DNA trajectories in plasma and serum separated by participant gender. Time courses of serum and plasma cf-mtDNA dynamics in stress and control visits: **(A,C)** serum cf-mtDNA, **(B,D)** plasma cf-mtDNA. Values in **(A,B)** represent averages of all measurements in both control and stress visits from participants a particular gender. Plots **(C,D)** show same data in (A,B) without averaging. Asterisks over symbols indicate a significant difference between average values from female and male participants. **(A-D)** Data shown as average ± standard error of the mean (SEM). Effect sizes and p-values from mixed-effects analyses (of log-transformed values). *p<0.05, **p<0.01, ***p<0.001, ****p<0.0001.

**Supplemental Figure 6.**
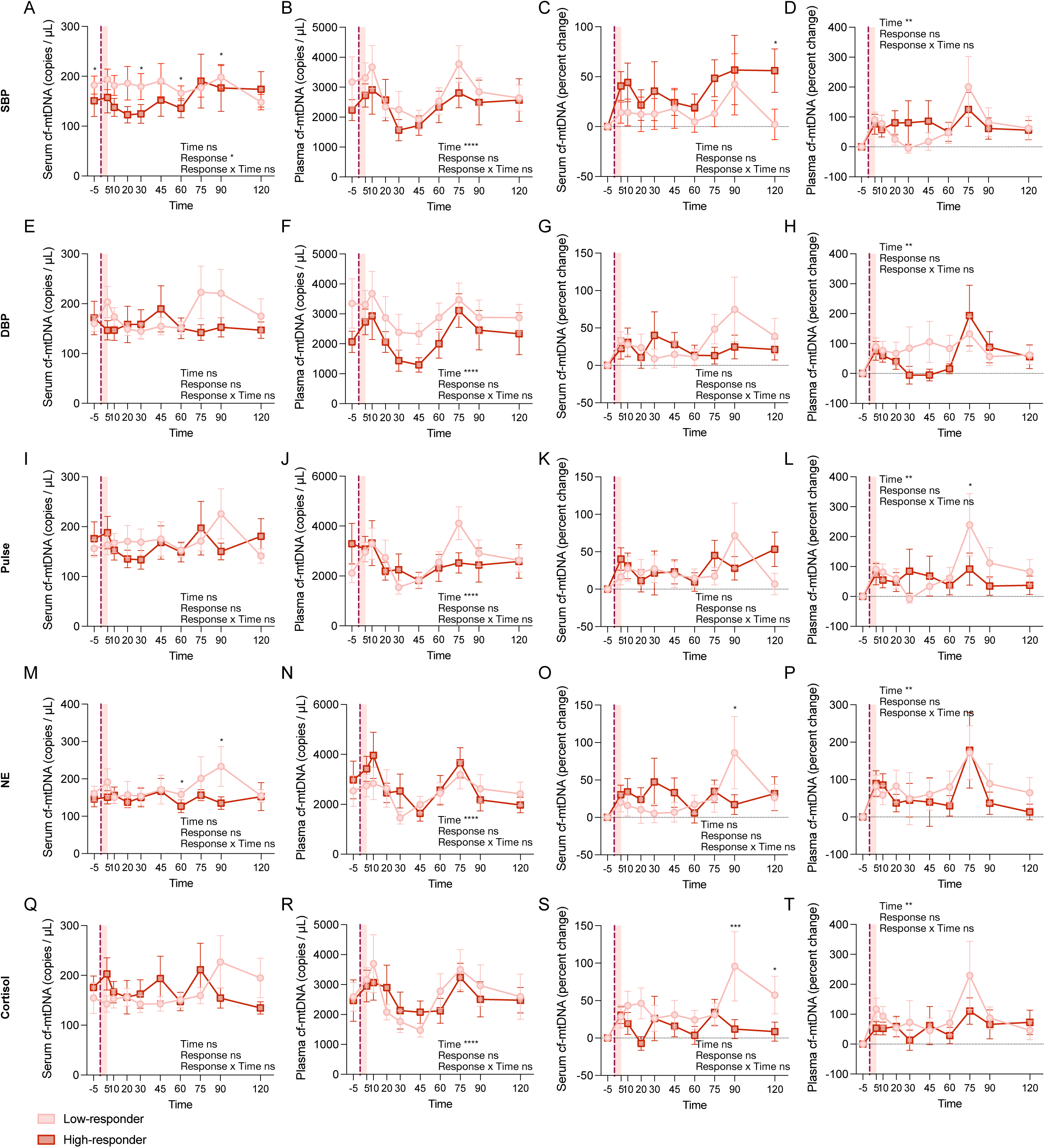
cf-mtDNA trajectories in response to psychological stress by participants cardiovascular and neuroendocrine responses. We evaluated whether participants cf-mtDNA response to the stress task differed by the magnitude of their cardiovascular and neuroendocrine stress response. Using a median split of reactivity to the stress task, we classified participants as low- and high-responders for individual measurements. Time course of cf-mtDNA during the stress visits for low and high systolic blood pressure (SBP) response for **(A)** serum cf-mtDNA copies of ND1 / µL and **(B)** plasma cf-mtDNA copies of ND1 / µL. **(C,D)** Same as in (A,B) for percent change from baseline. **(E-T)** Same as in (A-D) for **(E-H)** diastolic blood pressure (DBP), **(I-L)** pulse, **(M-P)** norepinephrine, **(Q-T)** cortisol. Asterisks over symbols indicate a significant difference between high and low responder values at the corresponding time point. Data shown as average ± standard error of the mean (SEM). (A-T) Effect sizes and p-values from mixed-effects analyses (A,B,E,F,I,J,M,N,Q,R log-transformed values). *p<0.05, **p<0.01, ***p<0.001, ****p<0.0001. Time windows used for calculating variable reactivities are defined in Supplemental Table 2.

**Supplemental Figure 7.**
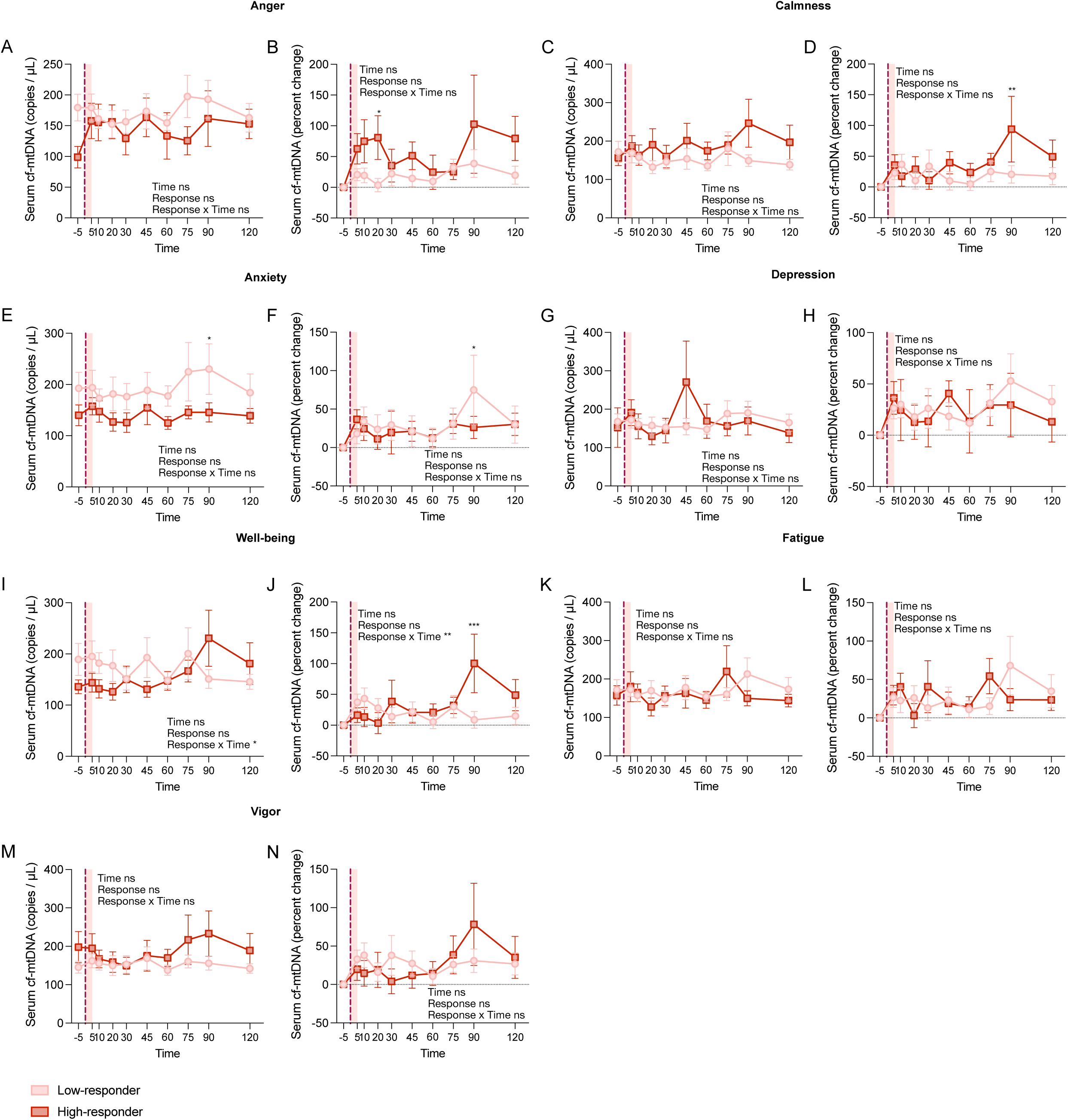
Serum cf-mtDNA trajectories in response to psychological stress by participants mood responses. We evaluated whether participants serum cf-mtDNA response to the stress task differed by the magnitude of their mood stress response. Using a median split of reactivity to the stress task, we classified participants as low- and high-responders for individual mood scores. Time course of serum cf-mtDNA during the stress visits for low and high anger responders as **(A)** copies of ND1 / µL and **(B)** percent change from baseline. **(C-N)** Same as in (A,B) for **(C,D)** calmness, **(E,F)** anxiety, **(G,H)** depression, **(I,J)** well-being, **(K,L)** fatigue, and **(M,N)** vigor. Asterisks over symbols indicate a significant difference between high and low responder values at the corresponding time point. Data shown as average ± standard error of the mean (SEM). (A-N) Effect sizes and p-values from mixed-effects analyses (A,C,E,G,I,K,M log-transformed values). *p<0.05, **p<0.01, ***p<0.001, ****p<0.0001. Time windows used for calculating variable reactivities are defined in Supplemental Table 2.

**Supplemental Figure 8.**
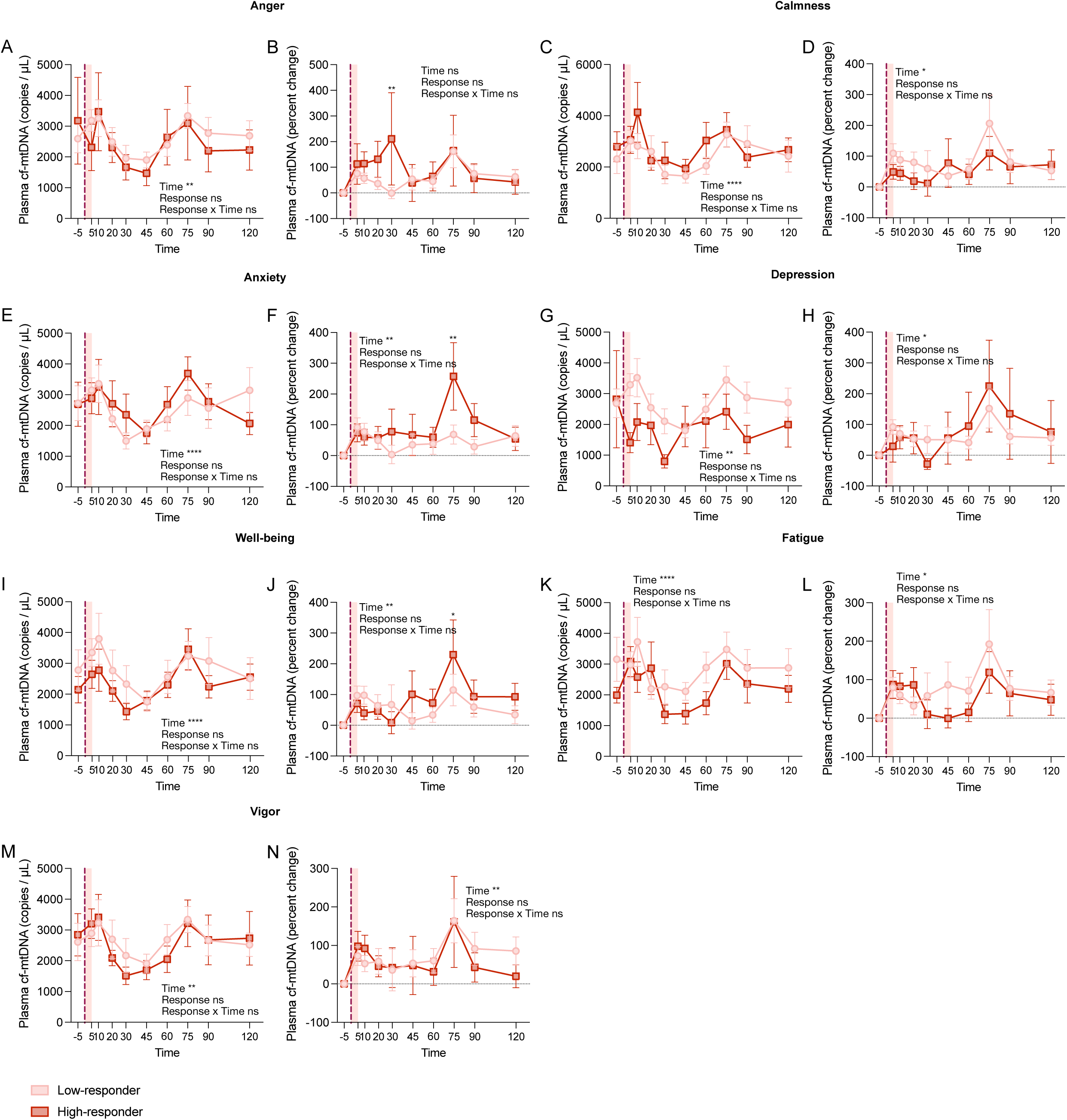
Plasma cf-mtDNA trajectories in response to psychological stress by participants mood responses. We evaluated whether participants plasma cf-mtDNA response to the stress task differed by the magnitude of their mood stress response. Using a median split of reactivity to the stress task, we classified participants as low- and high-responders for individual mood scores. Time course of plasma cf-mtDNA during the stress visits for low and high anger responders as **(A)** copies of ND1 / µL and **(B)** percent change from baseline. **(C-N)** Same as in (A,B) for **(C,D)** calmness, **(E,F)** anxiety, **(G,H)** depression, **(I,J)** well-being, **(K,L)** fatigue, and **(M,N)** vigor. Asterisks over symbols indicate a significant difference between high and low responder values at the corresponding time point. Data shown as average ± standard error of the mean (SEM). (A-N) Effect sizes and p-values from mixed-effects analyses (A,C,E,G,I,K,M log-transformed values). *p<0.05, **p<0.01, ***p<0.001, ****p<0.0001. Time windows used for calculating variable reactivities are defined in Supplemental Table 2.

